# Global reports of takotsubo (stress) cardiomyopathy following COVID-19 vaccination: a systematic review and meta-analysis

**DOI:** 10.1101/2022.04.16.22273937

**Authors:** Sirwan Khalid Ahmed, Mona Gamal Mohamed, Rawand Abdulrahman Essa, Eman Abdelaziz Ahmed Rashad Dabou, Salar Omar Abdulqadir, Rukhsar Muhammad Omar

## Abstract

Concerns have been raised recently about takotsubo cardiomyopathy (TCM) after receiving COVID-19 vaccines, particularly the messenger RNA (mRNA) vaccines. The goal of this study was to compile case reports to provide a comprehensive overview of takotsubo cardiomyopathy (TCM) associated with COVID-19 vaccines. A systematic literature search was conducted in PubMed, Scopus, Embase, Web of Science, and Google Scholar between 2020 and June 1, 2022. The study included individuals who developed cardiac takotsubo cardiomyopathy from receiving COVID-19 vaccinations. Ten studies, including 10 cases, participated in the current systematic review. The mean age was 61.8 years; 90% were female, while 10% were male. 80% of the patients received the mRNA COVID-19 vaccine, while 20% received other types. In addition, takotsubo cardiomyopathy (TCM) occurred in 50% of patients receiving the first dose and another 40% after the second dose of COVID-19 vaccines. Moreover, the mean number of days to the onset of symptoms was 2.62 days. All cases had an elevated troponin test and abnormal ECG findings. The left ventricular ejection fraction (LVEF) was lower than 50% in 90% of patients. In terms of the average length of hospital stay, 50% stayed for 10.2 days, and all cases recovered from their symptoms. In conclusion, takotsubo (stress) cardiomyopathy (TCM) complications associated with COVID-19 vaccination are rare but can be life-threatening. Chest pain should be considered an alarming symptom, especially in those who have received the first and second doses of the COVID-19 vaccine.

## Introduction

The coronavirus (COVID-19) pandemic, caused by the severe acute respiratory syndrome coronavirus-2 (SARS-CoV-2), has hurt millions of people worldwide. The FDA granted emergency approval to Pfizer-BioNTech (BNT162b2) and Moderna (mRNA-1273) COVID-19 vaccines in December 2020. SARS-CoV-2 vaccines are still the best hope for combating the global pandemic.

Takotsubo cardiomyopathy (TCM) is a type of heart disease frequently triggered by excessive physical and emotional stress. Although the pathophysiology of this condition is not fully understood, studies have suggested that it may be caused by the activation of central autonomic neurons expressing estrogen receptors. In addition, coronary vasospasm increases sensitivity to a surge in circulating catecholamines and metabolic dysfunction [1]. Catecholamine levels are higher in TCM than in STEMI [2,3]. As a result of the COVID-19 pandemic, TCM cases have risen from 1.5% to 7.8% [4]. TCM has already been linked to a ‘cytokine storm’ of interleukin 6 (IL-6) and tumor necrosis factor-alpha (TNFa), which causes hyper inflammation in COVID-19 patients [5,6]. Individuals who presented without COVID-19 infection or other notable risk factors may have been infected with TCM due to the psychological stress of living in a pandemic [1]. TCM is more likely to occur in people with a history of mental illness before the pandemic outbreak [5,7–9]. The mRNA1273 and BNT1626b2 (COVID-19 mRNA vaccines) induce innate immunity, cytotoxic and helper T cell responses, and, in particular, B cell responses [10]. Pain, erythema, swelling, fever, headache, and myalgia were reported as the most common systemic or local reactions [11]. However, acute myocardial infarction, pulmonary embolism, stroke, and venous thromboembolism are all possible complications of mRNA-based COVID-19 vaccines [12–14]. There have been reports of takotsubo cardiomyopathy following the COVID-19 vaccination, particularly following the messenger RNA (mRNA) vaccines [1,15–23].

We are aware of no systematic review that has been conducted specifically for COVID-19 vaccine induced TCM. Given the rarity of adverse takotsubo syndrome associated with COVID19 vaccination, most of the peer-reviewed papers are case reports. The goal of this study was to compile case reports to provide a comprehensive overview of takotsubo cardiomyopathy (TCM) associated with COVID-19 vaccines.

## Methods

### Review objectives

This study’s primary goal is to clarify the possibility of takotsubo cardiomyopathy (TCM) associated with COVID-19 vaccination. And to elaborate on the demographic and clinical characteristics of COVID-19 vaccinated individuals who develop takotsubo cardiomyopathy (TCM).

### Protocol and Registration

The review follows the PRISMA 2020 guidelines for reporting systematic reviews and meta-analyses [24]. The review protocol was recorded in the International Prospective Register of Systematic Reviews (PROSPERO) under the registration number CRD42022316515. Additionally, the AMSTAR-2 checklist was used to assess the quality of this review, which was found to be of high quality [25]. Ethical approval was not required for this review article.

### Search strategy

A comprehensive search of major online databases (PubMed, Scopus, Embase, Web of Science, and Google Scholar) was performed from 2020 until June 1, 2022, to discover all articles that had been published. The search strategy consisted of a combination of the following keywords: “COVID-19 vaccine” AND “takotsubo cardiomyopathy” OR “stress cardiomyopathy.” OR “takotsubo syndrome”. In addition, we checked the references of all relevant papers to ensure the search was complete.

### Eligibility criteria

1. All case series and case reports after the COVID-19 vaccine developed takotsubo cardiomyopathy (TCM) in humans, regardless of the type of Vaccine and dose, were included.
2. In this review, narrative and systematic reviews, as well as papers with insufficient data, were excluded. Furthermore, articles written in languages other than English were excluded.

### Data extraction and selection process

Every step of the data extraction process from the source was guided by PRISMA 2020. It was used to guide the process. Three independent authors (S.K.A., R.A.E., and M.G.M.) used the Rayyan website to screen abstracts and full-text articles based on inclusion and exclusion criteria [26]. Disagreements between the three independent authors were resolved by discussion. The data extraction process was carried out using predefined forms in Microsoft Office Excel. A total of the following information was extracted from each research study: author names, year of publication, age, gender, type of COVID-19 Vaccine, dose, troponin levels, days to symptom onset, symptoms, length of hospital stay/days, ECG, LVEF < 50% or LVEF > 50%, treatment, and their outcomes.

### Critical appraisal

We used the Joanna Briggs Institute’s critical appraisal tool for case reports to evaluate all included studies [27]. Three authors evaluated each article independently (S.K.A, M.G.M, and R.A.E). The paper evaluation disagreements were resolved through discussion or by the first author (S.K.A.). The results of our systematic review were evaluated using the AMSTAR 2 criteria [25]. The overall quality of our systematic review was rated “moderate” by the AMSTAR 2 tool.

### Data synthesis and analysis

The data from the articles included in this systematic review were extracted and pooled. This data included (author names, year of publication, age, gender, type of COVID-19 vaccine, dose, troponin levels, days to symptoms onset, symptoms, length of hospital stay/days, ECG, LVEF <50% or LVEF >50%, treatment, and their outcomes). We collected this information from the findings of eligible studies. Categorical data were expressed as proportions (%), and numerical data were expressed as mean ± standard deviation (SD). Statistical analysis will be performed using the Statistical Package for the Social Sciences (SPSS) 25.0 software program (SPSS Inc., Chicago, IL, USA). We pooled the results of the studies into one table.

## Results

### Selection of studies

The search was performed on significant databases (PubMed, Scopus, Embase, Web of Science, and Google Scholar) between 2020 and June 1, 2022, and yielded 2371 articles relevant to our search criteria. The references were then organized using a citation manager tool (Mendeley), and 459 articles were automatically eliminated because they were duplicates. Then, a final check was performed on 1912 articles to ensure that their titles, abstracts, and full texts were accurate. One thousand eight hundred ninety-seven articles were rejected because they did not meet the requirements to be included. Aside from that, 15 articles were submitted for retrieval, with five being dismissed because they did not meet our inclusion criteria. Finally, our systematic review included only ten articles (**Fig 1**). Case report details are shown in (**Table 1**).

**Table 1:**
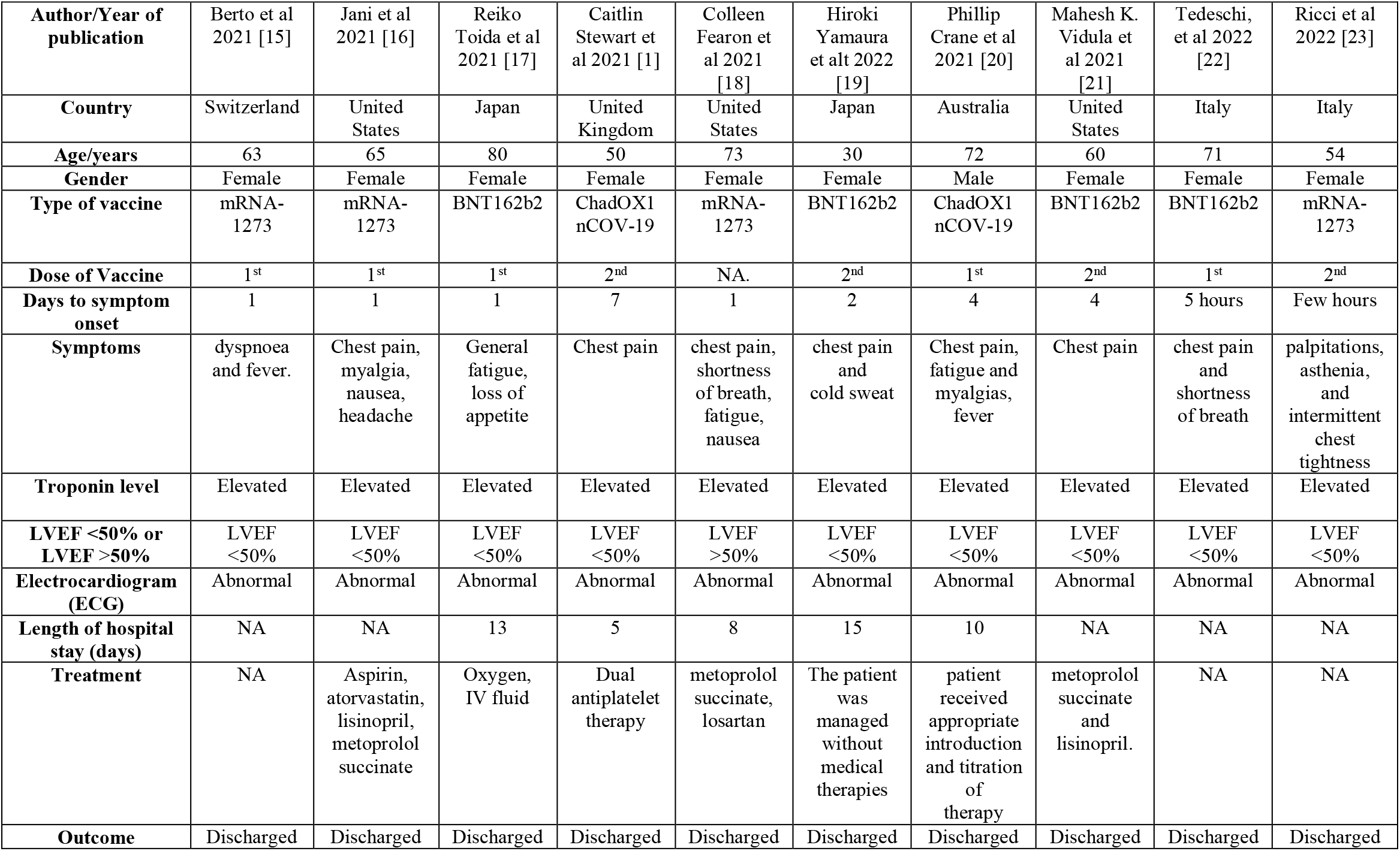
Characteristics and outcomes of patients with takotsubo cardiomyopathy related to COVID-19 Vaccine.

**Fig 1:**
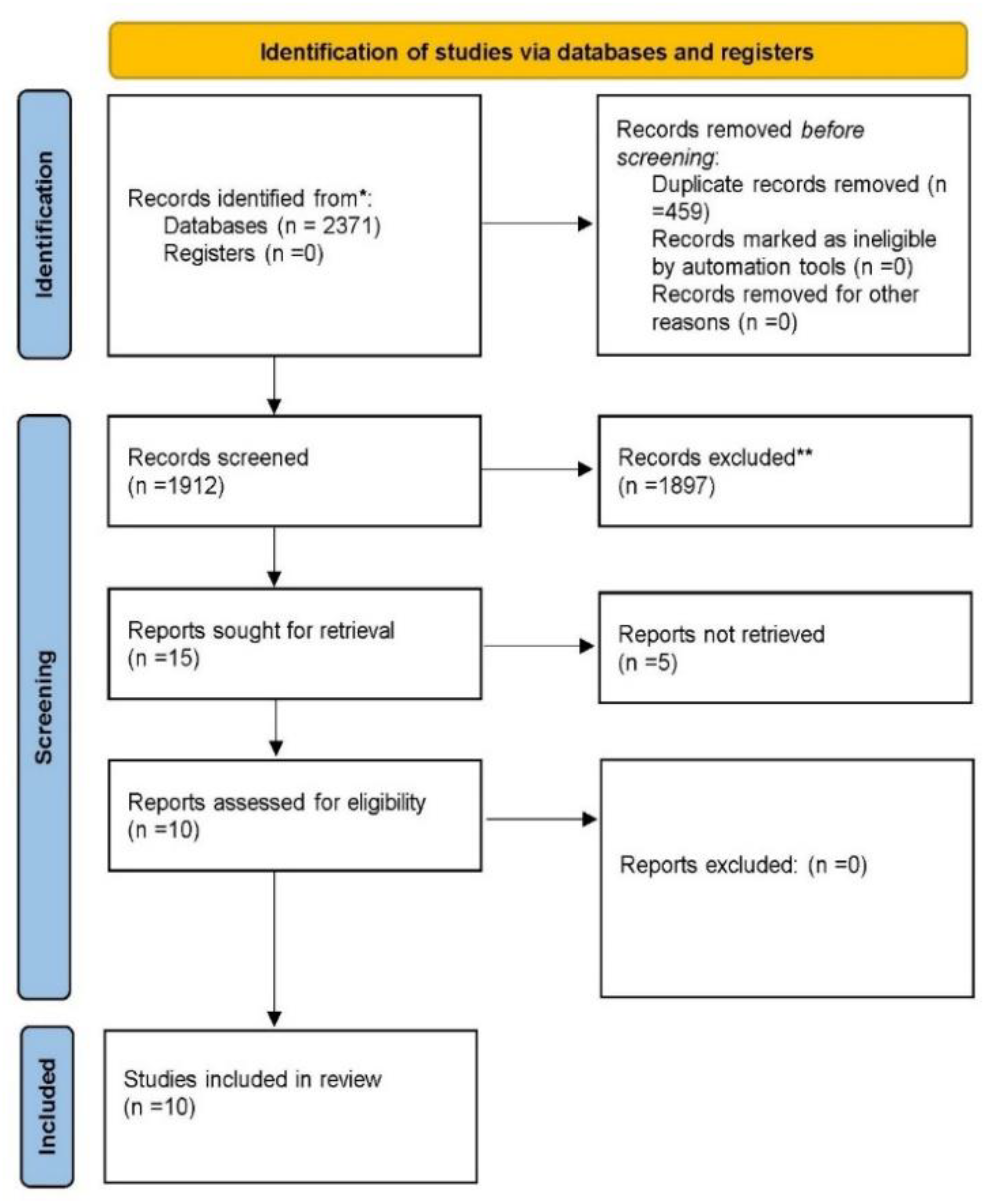
PRISMA flow diagram.

### Characteristics of the included studies

Table 2 shows an overview of the ten cases. The mean age and SD were 61.8 ±13.68 years; 90% were female, while 10% were male. 80% of the patients received the mRNA COVID-19 vaccine, either from Pfizer-BioNTech (BNT162b2) or Moderna COVID-19 Vaccine (mRNA-1273), while 20% of patients received other types, including AstraZeneca COVID-19 vaccine (ChAdOx1 nCoV-19). In addition, takotsubo cardiomyopathy occurred in 50% of patients receiving the first dose and another 40% after the second dose of COVID-19 vaccines. Moreover, the mean number of days to onset symptoms was 2.62 ± 2.19 days. All cases had an elevated troponin test and abnormal ECG findings. The left ventricular ejection fraction (LVEF) was lower than 50% in 90% of patients. Regarding the average length of hospital stay, 50% stayed for 10.20 ± 3.96 days, and all cases recovered from their symptoms.

**Table 2:** Summary of pooled data from included published research papers have been reported in the literature (n = 10)

| Variables | Results |
| --- | --- |
| Age (Mean ± SD) | 61.8 ± 13.68 years |
| Gender (n) % | Male – 1 (10%) |
|  | Female – 9 (90%) |
| Type of COVID-19 vaccine (n) % | Pfizer-BioNTech (BNT162b2) – 4 (40%) |
|  | Moderna COVID-19 Vaccine (mRNA-1273) - 4 (40%) |
|  | Oxford, AstraZeneca COVID-19 vaccine ChAdOx1 nCoV-19 – 2 (20%) |
| Dose (n) % | First dose – 5 (50%) |
|  | Second dose – 4 (40%) |
|  | None reported 1 (10%) |
| Days to symptom onset (Mean ± SD) | 2.62 ± 2.19 days |
| Troponin level (n) % | Elevated – 10 (100%) |
|  | Not elevated – 0 (0%) |
| Left ventricular ejection fraction LVEF (n) % | LVEF >50% - 9 (90%) |
|  | LVEF <50% - 1 (10%) |
| Electrocardiogram (ECG) (n) % | Abnormal – 10 (100%) |
|  | Normal – 0 (0%) |
| Length of hospital stay (days) (Mean ± SD) | 10.20 ± 3.96 days in 5 patients |
|  | Unknown in 5 patients |
| Outcome | Recovered – 10 (100%) |

## Discussion

COVID-19 vaccines protect the general public health and restrict viral propagation. Inactivated viral, DNA, mRNA, and protein-based vaccines are the four primary mechanisms that have been examined in this research for COVID-19 vaccines. DNA-based vaccinations use viral vectors to deliver the DNA coding for the SARS-CoV-2 spike protein into cells; mRNA vaccines typically deliver mRNA into cells via a lipid nanoparticle; protein vaccines use the spike proteins or their particles, and certain other vaccinations use inactivated viruses [44].

The COVID-19 pandemic has recorded the most extensive global vaccination program in history, with over 11.3 billion vaccination doses administered from December 2020 to December 2021. With increasing populations being vaccinated daily, we are seeing increased reports of associated side effects. The most common side effects are pain, swelling, and redness around the injection site. Systemic side effects like fever, tiredness, muscle pain, and headache can also happen to about one in four people. The females have noted a higher prevalence of these systematic effects [45]. Moreover, severe side effects have been reported and are under current monitoring: thrombocytopenic thrombosis, anaphylaxis, Bell’s palsy, myocarditis, and capillary leak syndrome [46,58,68].

In this systematic review, half of the cases were from the United States, most of the participants were female, and their mean age was 61.6. It was found that the participants received different types of COVID-19 vaccinations, including Pfizer-BioNTech (40 %), Moderna (mRNA-1273) (40%), and Oxford AstraZeneca ChAdOx1 nCoV-19 (20 %). More than half of the symptoms triggered by the first dose of the COVID-19 vaccine. Another important finding is that the symptoms started to happen after the vaccine dose, with a mean of 2.62 days. In line with the current results, Jani et al. (2021) mentioned that up to now, few cases of TCM have been reported after first or second dose administration of SARS-CoV-2 vaccines, even including different types of vaccines like mRNA-1273, BNT162b2, and ChAdOx1 nCoV-19 [47]. Another case report supported that a case of TCM after administration of the second dose of the DNA ChadOX1 nCOV-19 (AZD122) vaccination was reported, and this condition predominantly affects women (90%) at age 50 and above [48]. Also, Berto et al. (2021) wrote that a healthy 30-year-old Asian woman was brought to the emergency room with sudden chest pain and a cold sweat after receiving the second mRNA COVID-19 vaccine shot (Pfizer, New York City) 2 days prior [49].

Although the pathophysiology of TCM after COVID-19 vaccination is not well established, the proposed hypotheses are stunning changes in ischemia-induced myocardium. It could be secondary to microvessel or multi-vessel vasospasm and direct myocardial injury due to surge currents. Stress causes more release of catecholamine via excessive activation of sympathetic nervous system through the hypothalamic– pituitary-adrenal axis [50]. The limbic brain regions such as the amygdala, insula, anterior cingulate cortex, prefrontal cortex, and hippocampus have impaired neural networks during stress in TTS patients [51-54]. In a takotsubo rat model; epinephrin affinity switching from beta2-adrenoreceptors-Gs during low epinephrine amount to Gi when the amount of epinephrin is high causes acute apical cardiac depression, this process is protecting myocardium from toxicity during stress [55]. Scally et al., (2019) [6] has reported elevation of cytokines (serum interleukin-6, chemokine ligand 1, and classic CD14++CD16-) and infiltration of inflammatory myocardial macrophage in TTS patients. Naegele et al., (2016) [56] in his study found serious endothelial dysfunction in TTS patients. Another possible explanation is that the free-floating spike proteins may interact with angiotensin-converting enzyme 2. The imbalance between angiotensin II (overactivity) and angiotensin (deficiency) might play a role in the genesis of acute elevation in blood pressure, and this could be applied as a result of vaccination [62].

The diagnosis of takotsubo cardiomyopathy (TCM) was based on seven criteria put forward by the European Society of Cardiology [57]. The cardiac signs and symptoms, as well as electrocardiographic and laboratory findings, may be similar in patients who have TCM or myocarditis, which can make it very difficult to differentiate between the two conditions. Despite these difficulties, myocarditis is typically diagnosed with endomyocardial biopsy and cardiac magnetic resonance (CMR) [59]. Additionally, acute emotional or physical stressors induce increased levels of catecholamines and cortisol in the blood. These elevations mediate multiple pathways, including pericardial coronary artery spasms, microvascular dysfunction, and direct muscle cell injury, are all critical findings of transthoracic echocardiography [60,61].

In this systematic review, all cases had elevated troponin levels, the ejection fraction was above 50 %, and the electrocardiogram showed abnormal findings. According to the Mayo Clinic criteria, TCM has left ventricular hypokinesia, akinesia, or dyskinesia. These local wall dyskinesia are specific epicardial vasculature boundaries, with or without apex involvement. It is diagnosed when it expands beyond that. No evidence by angiography of obstructive coronary artery disease or acute plaque rupture, new ST elevation or T wave inversion, or mild cardiac troponin elevation. TCM is triggered by emotional, physical, or unknown causes [63,64].

These findings matched Toida et al. (2021), who reported that patients received the first dose of the Pfizer-BioNTech COVID-19 vaccine. The day after the vaccination, she developed general fatigue and a loss of appetite. A physical examination revealed a systolic murmur of Levine 2/6 at the second left sternal border, clear lung sounds, and no leg edema. On admission, electrocardiography (ECG) revealed atrial fibrillation with a normal axis, negative T-waves in I, aVL, V3-6 leads, and a prolonged QTc interval of 495 ms [65].

In our review, more than half of the patients stayed in the hospital with a mean ± SD of 10.20 ± 3.96 days, and all recovered without complications. An outcomes study assessing the trends of hospitalized patients with takotsubo cardiomyopathy demonstrated an in-hospital mortality rate of 1.3%, a hospital discharge rate to the home of 73.6%, and 1-year mortality of 6.9% [66]. In addition, Đenić et al. (2022) reported that the patient was clinically improved and had no evidence of high-risk features on ECG or TTE [67]. However, these findings do not support the previous study by Toida et al. (2021), who showed the case report regarding takotsubo cardiomyopathy after the COVID-19 vaccine and found the patient’s length of hospital stay was about five days [65].

## Limitation

We recognize that our systematic review has some limitations, which include only involving case reports and case series due to the limited numbers of original studies published on the takotsubo cardiomyopathy after taking COVID-19 vaccines, so the results should be interpreted with some caution. Moreover, we do not have any data about the previous history of COVID-19, which may affect the results since COVID-19 can also cause cardiac complications.

## Conclusion

Takotsubo cardiomyopathy (TCM) complications associated with COVID-19 vaccination are rare but can be life-threatening. Chest pain should be considered an alarming symptom, especially in those who have received a second dose of the vaccine in the last three days. For diagnosis, CK-MB and troponin are better biomarkers to confirm myocarditis than CRP, ESR, and NT-proBNP. All of the cases had wholly recovered without any irreversible cardiomyopathy changes.

## Data Availability

All relevant data are within the manuscript and its supporting information files.

## Conflicts of interest

There is no conflict to be declared.

## Funding

This research did not receive any specific grant from the public, commercial, or not-for-profit funding agencies.

## Author Agreement Statement

We declare that this manuscript is original, has not been published before, and is not currently being considered for publication elsewhere. We confirm that the manuscript has been read and approved by all named authors and that there are no other persons who satisfied the criteria for authorship but are not listed. We confirm that all have agreed with the order of authors listed in our manuscript. We understand that the Corresponding Author is the sole contact for the Editorial process. He is responsible for communicating with the other authors about progress, submissions of revisions, and final approval of proofs.

## Data availability Statement

All relevant data are within the manuscript and its supporting information files.

## Authors’ contributions

Conception and design SKA acquisition of data SKA, RAE analysis and interpretation of data SKA, RAE, MGM, EEA, drafting of the manuscript SKA, RAE, MGM, EAA critical revision of the manuscript for effective intellectual content statistical analysis SKA, SAO and RMO, administrative SKA, RAE, MGM, EAA technical SKA, supervision SKA and all authors approving the final draft.

## Provenance and peer review

Not commissioned, externally peer-reviewed

## Acknowledgments

Not applicable

## Notes

### Competing Interest Statement

The authors have declared no competing interest.

### Funding Statement

This research did not receive any specific grant from funding agencies in the public, commercial, or not-for-profit sectors

### Summary of Updates

The discussion section was revised again

